# Uveal Melanoma: A Preliminary Analysis of Incidence in Irkutsk Oblast, 2023–2025, and Potential Causes of Elevated Rates Compared to Global and National Data

**DOI:** 10.64898/2026.07.29.26359176

**Authors:** Yuri Senkin, Olga Senkina

**Affiliations:** Irkutsk Regional Oncocentre, ul. Frunze,32, Irkutsk, 664035, Russia

## Abstract

**Objective:** To analyze the incidence dynamics of uveal melanoma (UM) in Irkutsk Oblast over the period 2023–2025, assess the structure of newly diagnosed cases by territory and stage, and compare the outcomes of anticancer care with those of other constituent entities of the Russian Federation that lead in registered prevalence of malignant neoplasms (MN) of the eye and adnexa.

**Materials and Methods:** Data from the cancer registry of the Irkutsk Regional Oncology Dispensary (IROD) for 2023–2025 and information from the state reference book "The State of Cancer Care for the Population of Russia in 2025" (P.A. Herzen Moscow Oncology Research Institute) were used. The analysis included cases of MN of the eye and adnexa (ICD-10 code C69) of uveal localization. Incidence rates per 100,000 population, stage distribution, proportion of morphological verification, mortality, and the cohort of patients under follow-up were calculated.

**Results:** Over three years, 112 new cases of UM were registered in Irkutsk Oblast (37, 36, and 39, respectively), corresponding to a crude incidence rate of approximately 1.6 per 100,000 population per year. In 2025, the region ranked 3rd in the Russian Federation for the number of newly registered patients with MN of the eye (45 individuals) and 1st for the registered prevalence rate (15.6 per 100,000). The highest proportion of stage I disease (58.3 %) and one of the lowest mortality rates (1.9 %) were observed.

**Conclusions:** Irkutsk Oblast is characterized by a consistently high incidence of UM, accompanied by a favorable stage distribution and low mortality, reflecting both the biological and geographic features of the region and an adequate level of organization of ophthalmic oncology care.

## Introduction

Uveal melanoma (UM) is the most common primary intraocular malignancy in adults and ranks second among melanocytic neoplasms after cutaneous melanoma [1]. The tumour arises from melanocytes of the uveal tract—the iris, ciliary body, and choroid—with choroidal localisation accounting for the majority of cases. Despite its relative rarity, UM has a high metastatic potential: haematogenous dissemination, predominantly to the liver, determines an unfavourable long-term prognosis even when local control of the primary tumour is achieved [2]. Worldwide, the incidence of UM is estimated at 5–8 cases per million population per year (approximately 0.5–0.8 per 100,000), with a pronounced geographical gradient. The highest rates are reported in Northern European countries—Norway, Denmark, and Sweden—where incidence reaches 8–10 per million [3, 4]. According to the large European EUROCARE study, the average European rate is 4.4 per million, ranging from 2 cases (Spain, southern Italy) to 8 cases (Norway, Denmark) per million [3]. This gradient has traditionally been attributed to the prevalence of light-skinned phenotypes in northern populations and an inverse relationship between incidence and solar insolation intensity. It is important to emphasise that this latitudinal gradient fundamentally distinguishes uveal melanoma from cutaneous melanoma, which exhibits a direct association with cumulative ultraviolet exposure; the pathogenic role of sunlight in UM development remains debated, with constitutional phenotypic factors considered the predominant drivers. According to large population-based studies, UM incidence rates have remained relatively stable over recent decades without marked increases, in contrast to cutaneous melanoma [5].

In the Russian Federation, systematic population-based data on UM incidence are limited, and in official statistics the tumour is recorded within the broader category "malignant neoplasms of the eye and adnexa" (ICD-10 code C69). The most comprehensive national data come from Moscow: according to E.E. Grishin and co-authors, the UM incidence rate was approximately 0.9–1.07 per 100,000 adult population over the observation period 1977–2012 [6, 7]. The mean age at diagnosis ranges from 60 to 67 years, with a peak in the 70–79 age group. Established risk factors include fair phenotype (blue or green eyes, light skin), residence at higher geographical latitudes, oculodermal melanocytosis, as well as tumour molecular-genetic features—somatic mutations in *GNAQ/GNA11*, found in about 90 % of cases, and germline or somatic *BAP1* mutations, which are associated with a high risk of metastasis [1, 5]. The variability of regional incidence rates within Russia has been insufficiently studied, complicating the planning of specialised ophthalmic oncology care.

The clinical course and prognosis of UM are determined by several factors, among which the most critical are primary tumour size, localisation, histological type (spindle-cell, epithelioid-cell, or mixed), presence of extrascleral extension, and the molecular-genetic profile. It has been established that the risk of distant metastasis increases almost linearly with increasing maximal basal diameter and tumour thickness: in an analysis of over 8,000 cases, each additional millimetre of tumour thickness was associated with an increment in cumulative metastatic risk [2]. This underscores the paramount importance of early diagnosis: detection at a small size enables eye-preserving treatments—brachytherapy, transpupillary thermotherapy, stereotactic radiotherapy—and substantially improves both functional and survival outcomes [1, 7]. Despite advances in local control of the primary lesion, long-term survival rates for UM have changed little over recent decades, owing to early subclinical haematogenous dissemination and the absence of highly effective systemic therapy for metastatic disease [5]. These features highlight that the main reserve for improving outcomes remains early tumour detection; consequently, studying the epidemiological picture and the pattern of case detection at the population level has practical implications for the organisation of ophthalmic oncology services.

Of particular interest is Irkutsk Oblast, which, according to the state reference book *"The State of Cancer Care for the Population of Russia in 2025"*, ranks among the leading subjects of the Russian Federation in the number of newly registered patients with malignant neoplasms of the eye and adnexa, and also leads in the registered prevalence of this pathology [8]. The region is located in the southern part of Eastern Siberia at approximately 52° N latitude, corresponding to high latitudes with relatively low annual insolation; a considerable proportion of the population exhibits a light-skinned phenotype. The combination of these factors makes Irkutsk Oblast a suitable model for studying regional UM epidemiology and evaluating the outcomes of specialised care. At the same time, data on regional UM epidemiology in the Russian Federation remain fragmentary: most domestic publications are based on materials from individual specialised centres and do not reflect population differences between constituent entities. In this context, comparing regional cancer registry data with the national reference book not only allows characterisation of the epidemiological situation in a specific region but also enables assessment of registration completeness and diagnostic quality in an interregional comparison.

This study is devoted to analysing the incidence dynamics of UM in Irkutsk Oblast over 2023–2025, examining the territorial and stage-related structure of newly diagnosed cases, and comparing the main cancer care indicators with those of leading regions—Moscow, Moscow Oblast, and Saint Petersburg. The working hypothesis is that the high incidence and prevalence rates of UM in Irkutsk Oblast reflect a combined effect of biological-geographic determinants and organisational factors that ensure complete case detection and registration.

From a clinical and morphological perspective, UM represents a heterogeneous group of tumours differing by localisation (iris, ciliary body, choroid), histological architecture, and molecular-genetic characteristics. Iris melanomas are usually detected earlier owing to their accessibility to examination and have a more favourable prognosis, whereas ciliary-body and choroidal tumours are often asymptomatic for a long time. Current understanding of UM molecular pathogenesis links tumour initiation to activating mutations in *GNAQ* and *GNA11*, found in approximately 90 % of cases, and progression and metastatic potential to inactivation of the tumour suppressor gene *BAP1*, as well as mutations in *SF3B1* and *EIF1AX* [1]. Monosomy of chromosome 3 is considered one of the strongest predictors of metastasis. Despite progress in understanding tumour biology, molecular-genetic profiling in routine Russian ophthalmic oncology practice remains limited, further increasing the importance of clinical-epidemiological data for characterising the regional situation. Diagnosis of UM in modern clinical practice relies primarily on non-invasive imaging methods. Key modalities include ophthalmoscopy under pharmacological mydriasis, B-mode ocular ultrasound to assess tumour dimensions and acoustic structure, as well as optical coherence tomography and fluorescein angiography. Together, these methods provide high accuracy of clinical diagnosis; consequently, histological verification prior to treatment is performed much less frequently for intraocular tumours than for neoplasms at most other sites [1]. This is a crucial consideration for correct interpretation of the morphological verification rate in interregional comparisons, and it explains why a relatively low proportion of pathological confirmation in UM should not be regarded as a sign of inadequate diagnostic quality. The modern therapeutic armamentarium includes eye-preserving approaches—brachytherapy with ophthalmic applicators, transpupillary thermotherapy, stereotactic radiotherapy, and proton therapy—as well as ablative surgery (enucleation) for extensive involvement. The choice of modality is determined by tumour size and localisation, visual function, and the patient’s general condition; the key prerequisite for applying conservative techniques remains timely detection at small to medium size [1, 7].

The relevance of regional epidemiological studies of UM is determined by several considerations. First, the rarity of the tumour and its inclusion within the aggregated ICD-10 category C69 mean that national statistics do not reflect the specifics of uveal localisation, and population differences across the country remain unaddressed. Second, planning specialised ophthalmic oncology care—including the placement of brachytherapy centres and the training of relevant specialists—requires objective data on the territorial distribution of incidence. Third, identifying regions with anomalously high incidence provides an opportunity to study environmental and population-genetic determinants of tumour development, which has implications for fundamental understanding of its aetiology [5]. Collectively, these arguments justify the need for systematic analysis of regional cancer registry data and their comparison with official national statistics, which constitutes the subject of the present study.

## Materials and Methods

A retrospective analysis was conducted on data regarding newly diagnosed cases of UM among the population of Irkutsk Oblast from 1 January 2023 through 31 December 2025. The primary data source was the population-based cancer registry of the Irkutsk Regional Oncology Dispensary (IROD), which is the region’s leading specialised institution where diagnosis and treatment of patients with ophthalmic oncology pathology are centralised. The registry accumulates information on all first-ever diagnoses of malignant neoplasms in residents of the region, regardless of the site of initial presentation.

The inclusion criterion was a first-ever diagnosis of a malignant neoplasm of the eye and adnexa (ICD-10 code C69) with confirmed uveal localisation—involvement of the uveal tract (iris, ciliary body, or choroid), corresponding to subcategories C69.3 (choroid) and C69.4 (ciliary body) upon morphological and/or clinical-instrumental verification of melanoma. Neoplasms of other localisations within the C69 rubric (conjunctiva, cornea, lacrimal gland) and secondary (metastatic) lesions of the uveal tract were excluded from the analysis.

For comparative analysis and to characterise the position of Irkutsk Oblast among the constituent entities of the Russian Federation, data from the official state reference book *The State of Cancer Care for the Population of Russia in 2025"* were used. This reference book was prepared by the P.A. Herzen Moscow Oncology Research Institute—a branch of the National Medical Research Centre of Radiology of the Ministry of Health of the Russian Federation—under the editorship of A.D. Kaprin, V.V. Starinsky, A.O. Shakhzadova, and A.D. Eremeeva [8]. Information was extracted from the reference book for rubric C69 for the Russian Federation as a whole and for selected constituent entities— Moscow, Moscow Oblast, Saint Petersburg, and Irkutsk Oblast—allowing regional data to be placed in a national context.

The following indicators were analysed: absolute number of newly detected cases by year and their distribution across the administrative territories of the region (Irkutsk city, Angarsk, Bratsk, Ust-Ilimsk, and other districts). The crude (non-standardised) incidence rate was calculated as the ratio of new cases per year to the average annual population of the region, expressed per 100,000 population; the estimated population of Irkutsk Oblast in the range of 2.3–2.4 million people was used for the calculations. The distribution of cases by tumour stage (I–IV) according to the clinical classification, the proportion of morphologically verified diagnoses, the mortality rate, and the cohort of patients under follow-up, including the proportion of persons under observation for 5 years or more, were assessed. The registered prevalence rate (patients under follow-up at the end of the year) was calculated per 100,000 population.

Data are presented using descriptive statistics as absolute numbers, relative frequencies (%), and crude rates per 100,000 population. Owing to the limited observation period and the nature of the aggregated source data, age standardisation of rates and statistical significance testing were not performed; comparisons are descriptive in nature. For cases from 2025, the stage distribution is presented according to the state reference book [8], because at the time of analysis, the regional registry data for that year had not been detailed by stage.

For interregional comparisons, the following comparison regions were deliberately selected: the constituent entities of the Russian Federation with the highest absolute numbers of newly registered patients—Moscow, Moscow Oblast, and Saint Petersburg—representing the country’s largest urban agglomerations with well-developed specialised ophthalmic oncology services. This selection allows comparison of Irkutsk Oblast indicators not with an "average" region but with territories that have the maximum diagnostic capabilities, making the comparison as conservative as possible with respect to evaluating detection completeness. All data used are anonymised aggregated statistical indicators and do not contain personal information, in accordance with the ethical requirements for retrospective epidemiological studies.

The completeness and quality of the source data are ensured by the organisation of the region’s oncology service. According to the current procedure for medical care in the field of oncology, all first-ever diagnoses of malignant neoplasms are subject to mandatory registration in the population-based cancer registry, and patients with intraocular tumours are concentrated at the IROD as the leading specialised institution with capabilities for ophthalmic oncology diagnosis and treatment. This centralisation minimises the risk of incomplete registration and duplication of records—issues commonly encountered with pathologies where diagnosis is dispersed across many facilities. Verification of uveal localisation was based on ophthalmoscopy, ocular ultrasound, and, when available, pathological reports; doubtful or unspecified observations were excluded from the analysis. Data extraction from the registry was performed using a complete enumeration method for the entire observation period without sampling procedures, ensuring representativeness of the obtained information for the entire patient population of the region.

## Results

### Incidence dynamics of uveal melanoma in 2023–2025

Over the three-year observation period, 112 new cases of UM were registered in Irkutsk Oblast. The annual distribution was as follows: 37 cases in 2023, 36 cases in 2024, and 39 cases in 2025. Thus, the average annual number of newly detected cases remained within a narrow range of 37–39 observations, indicating stable incidence without marked fluctuations. With the average annual population of the region being approximately 2.3–2.4 million, the crude incidence rate was about 1.6 cases per 100,000 population per year. This value substantially exceeds the average European level (0.44 per 100,000) and is comparable to or higher than the rates previously reported for Moscow (0.9–1.07 per 100,000 adult population) [3, 6]. The dynamics of absolute numbers and crude rates are presented in Table 1.

**Table 1:** Dynamics of Uveal Melanoma incidence in Irkutsk Oblast, 2023–2025.

| **Indicator** | **2023** | **2024** | **2025** |
| --- | --- | --- | --- |
| Number of new cases, abs. | 37 | 36 | 39 |
| Population, million (estimate) | 2,4 | 2,4 | 2,3 |
| Incidence rate per 100,000 population | ≈1,5 | ≈1,5 | ≈1,7 |
| **Total for 3 years, abs.** | **112** |  |  |

*Note. The incidence rate is calculated as a "rough" (non-standardized) intensive indicator. Source: oncological registry of GBUZ IOOD*

The stability of absolute numbers over the three years points to an established epidemiological level of incidence in the region rather than a random spike in detectability. The slight increase in 2025 (to 39 cases) falls within the expected interannual fluctuations for a rare tumour and cannot be interpreted as a reliable trend given the limited observation period. It should be emphasised that the obtained incidence rate (≈ 1.6 per 100,000) is calculated for the entire population, including children, whereas UM is virtually absent in individuals under 20 years of age and is concentrated in age groups over 50 years. Consequently, the true incidence rate among the adult population—which accounts for the vast majority of cases—is naturally higher than the crude rate calculated for the total population, even more strongly highlighting the region’s leading position. Comparison of the absolute number of new cases in the region (37–39 per year) with the data from the state reference book (45 registered in 2025 for the entire C69 rubric) is consistent with UM accounting for the predominant share of malignant eye neoplasms in the adult population.

Analysis of absolute numbers in comparison with national data underscores the scale of the issue: although the population of Irkutsk Oblast constitutes about 1.6 % of the Russian Federation’s population, the region accounts for a considerably larger proportion of newly registered patients with eye neoplasms (45 out of 1379, i.e. approximately 3.3 % of the national total). This two-fold excess of the share in the incidence structure over the share in the population quantitatively confirms the region’s leading position in the incidence and prevalence of UM. It should be noted that the absolute number of cases according to the regional registry (39 in 2025) and the state reference book (45) are close; the discrepancy is explained by the reference book covering the entire C69 rubric, including non-uveal localisations, which further confirms that UM predominates in the structure of malignant eye neoplasms among the adult population of the region.

### Territorial structure of newly detected cases

Analysis of the territorial distribution of detected cases reflects both the population settlement pattern of the region and the organisation of primary ophthalmic care. In 2023, of 37 cases, 16 were recorded in Irkutsk city, 3 in Angarsk, 6 in Bratsk, and 12 in other districts of the region. In 2024, of 36 cases, 15 were in Irkutsk city, 3 in Angarsk, 2 in Bratsk, 3 in Ust-Ilimsk, and 13 in other districts. In 2025, of 39 cases, 18 were in Irkutsk city, 6 in Angarsk, 2 in Bratsk, and 13 in other districts. Thus, the regional capital consistently accounts for the largest proportion of detected cases (averaging about 43–46 %), which is expected given the concentration of population and the specialised ophthalmic service. At the same time, a substantial proportion of cases (on average more than one-third) come from districts outside the major cities, underscoring the need to maintain diagnostic awareness at the primary care level in remote areas. Detailed territorial distribution is presented in Table 2.

**Table 2:**
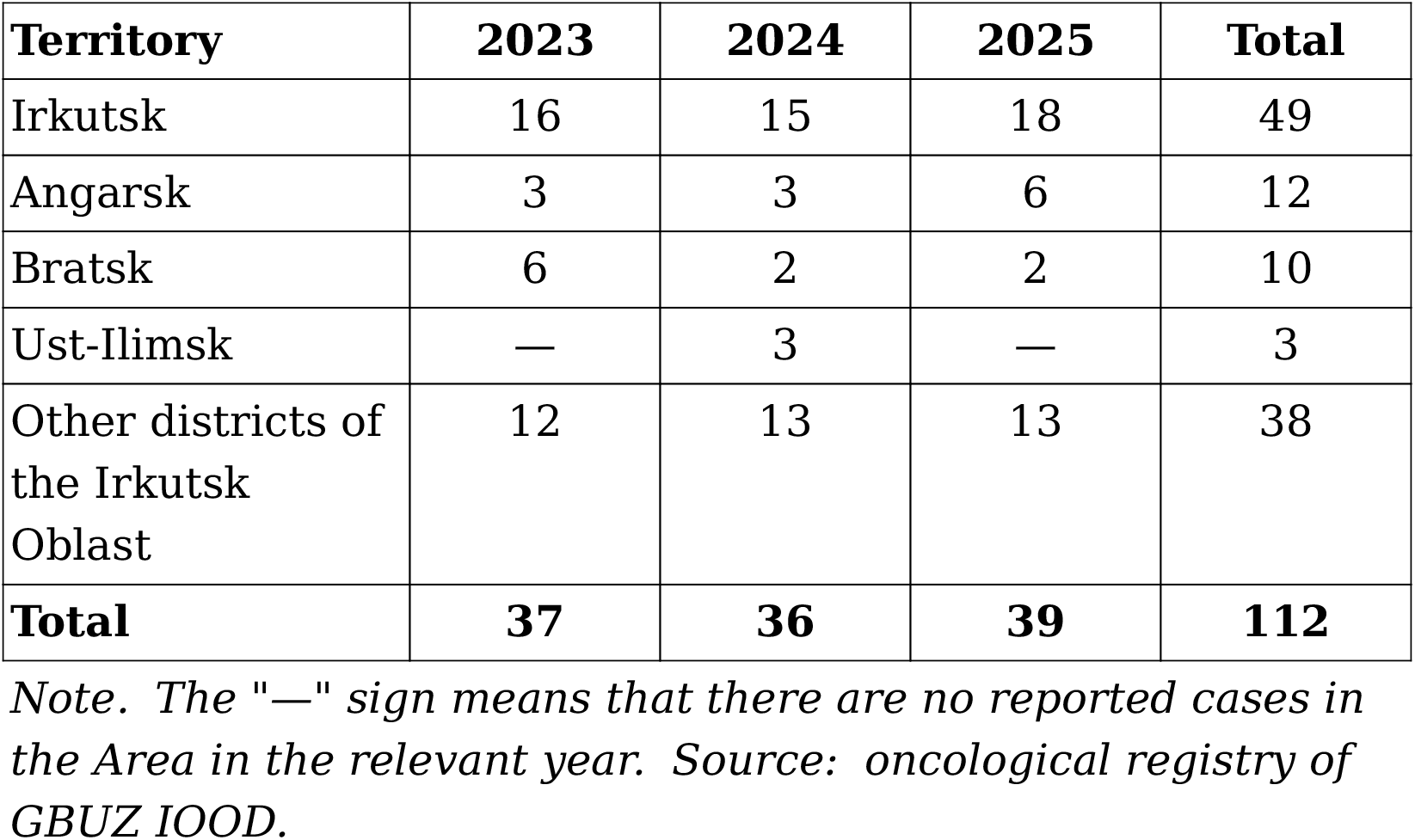
Territorial distribution of newly diagnosed cases of uveal melanoma in the Irkutsk region, 2023–2025 (abs.)

| **Territory** | **2023** | **2024** | **2025** | **Total** |
| --- | --- | --- | --- | --- |
| Irkutsk | 16 | 15 | 18 | 49 |
| Angarsk | 3 | 3 | 6 | 12 |
| Bratsk | 6 | 2 | 2 | 10 |
| Ust-Ilimsk | — | 3 | — | 3 |
| Other districts of the Irkutsk Oblast | 12 | 13 | 13 | 38 |
| **Total** | **37** | **36** | **39** | **112** |

*Note. The "—" sign means that there are no reported cases in the Area in the relevant year. Source: oncological registry of GBUZ IOOD.*

Over the three-year period, Irkutsk city accounted for a total of 49 cases (43.8 % of all observations), other districts of the region for 38 cases (33.9 %), Angarsk for 12 (10.7 %), Bratsk for 10 (8.9 %), and Ust-Ilimsk for 3 (2.7 %). Thus, about three-quarters of all cases are concentrated in the regional centre and rural districts, reflecting two opposing trends: centralisation of detection in the centre with a well-developed specialised service, and a persistent significant proportion of patients from remote areas who are referred to the IROD for diagnostic verification and treatment. The relatively low share of large industrial cities (Angarsk, Bratsk, Ust-Ilimsk) may be related both to smaller populations in older age groups and to specific patient referral pathways, and requires further analysis considering the age structure of the population in these territories. The considerable contribution of rural districts (on average over one-third of cases) indicates that diagnostic awareness at the primary care level in remote areas of the region is at an adequate level, ensuring patient referral to the specialised facility.

### Stage distribution and comparison with regions of the Russian Federation

The distribution of newly detected cases by tumour stage demonstrates a predominance of early stages throughout the observation period according to the regional registry data. In 2023, of 37 cases, stage I was established in 22 patients, stage II in 8, stage III in 6, and stage IV in 1. In 2024, of 36 cases, stage I accounted for 20 observations, stage II for 8, stage III for 6, and stage IV for 2. For 2025, the stage distribution is presented according to the state reference book [8]: the proportion of stage I was 58.3 %, stage II – 16.7 %, stage III – 22.9 %, stage IV – 2.1 %, with no cases of unknown stage. The annual stage dynamics are presented in Table 3.

**Table 3:**
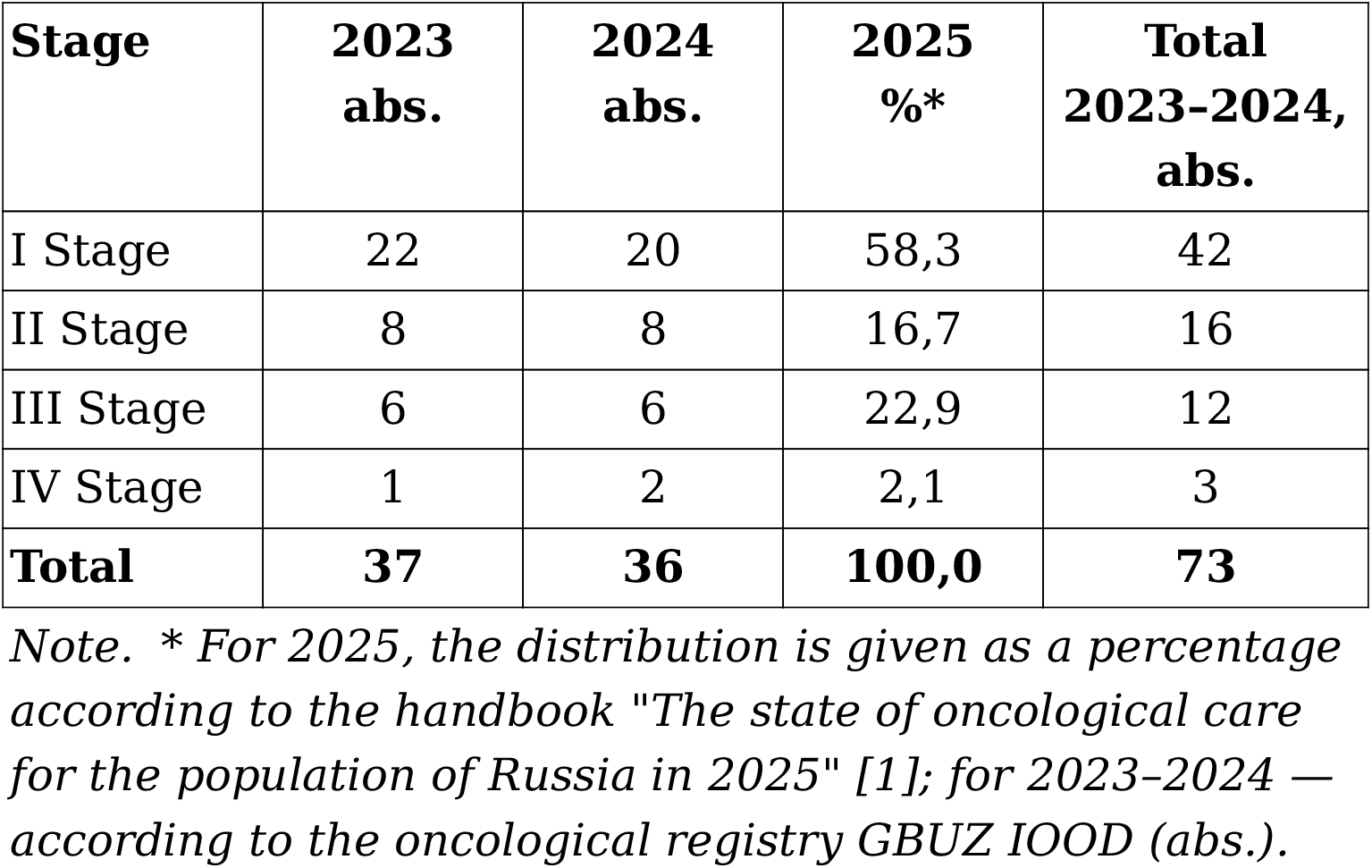
Distribution of newly diagnosed cases of uveal melanoma by stage in the Irkutsk region, 2023–2025.

Comparison of the stage distribution at detection in 2025 with the leading regions by registered prevalence reveals a clear advantage for Irkutsk Oblast in terms of early diagnosis. The proportion of stage I in the region (58.3 %) substantially exceeds the corresponding figures for Moscow (32.6 %), Moscow Oblast (25.3 %), and Saint Petersburg (28.3 %) [8]. At the same time, Irkutsk Oblast has no cases of unknown stage (0 %), whereas in Moscow Oblast the proportion of such observations reaches 34.7 %, and in Saint Petersburg – 28.3 %, which complicates the correct interpretation of comparative data for these regions. The proportion of stage II in Irkutsk Oblast, conversely, was lower than in the comparator regions (16.7 % vs. 47.4 % in Moscow, 28.0 % in Moscow Oblast, and 35.8 % in Saint Petersburg), which is largely explained by differences in staging approaches and completeness of recording. Comparative indicators of cancer care under rubric C69 are presented in Table 4.

**Table 4:** Comparative characteristics of indicators of oncolog­ ical care for neoplasms of the eye and appendage (C69) in cer­ tain regions of the Russian Federation, 2025.

| **Indicator** | **Irkutsk Oblast** | **The city of Moscow** | **Moscow Oblast** | **The city of St. Petersburg** |
| --- | --- | --- | --- | --- |
| Registered in 2025, abs. | 45 | 93 | 75 | 51 |
| Prevalence on the register, per 100 000 | 15,6 | 11,7 | 7,4 | 11,9 |
| Registered for 5 years or more, % | 55,5 | 72,1 | 52,8 | 68,5 |
| The Mortality rate, % | **1,9** | 2,9 | 1,4 | 3,0 |
| Morphological verification, % | 56,3 | 53,7 | 100 | 94,3 |
| I Stage, % | **58,3** | 32,6 | 25,3 | 28,3 |
| II Stage, % | 16,7 | 47,4 | 28,0 | 35,8 |
| III Stage, % | 22,9 | 15,8 | 10,7 | 7,5 |
| IV Stage, % | 2,1 | 0 | 1,3 | 0 |
| Unidentified stage, % | **0** | 4,2 | 34,7 | 28,3 |

*Note. Source: handbook "The state of oncological care for the Russian population in 2025" [8]. The most favorable values for the region are shown in bold.*

The set of indicators presented in Table 4 forms an internally consistent picture, characterising Irkutsk Oblast as a region with a high prevalence of UM accompanied by a favourable detection structure. The highest registered prevalence rate among the compared regions (15.6 per 100,000) is combined with the highest proportion of stage I (58.3 %), one of the lowest mortality rates (1.9 %), and a complete absence of cases with unknown stage [8]. The combination of high prevalence with low mortality is a characteristic sign of favourable survival and accumulation of the cohort of patients under follow-up, because with early detection and effective local control, most patients remain under dispensary observation for prolonged periods. The high proportion of cases with unknown stage in Moscow Oblast (34.7 %) and Saint Petersburg (28.3 %) is noteworthy, as it limits the validity of direct comparisons of stage structure and may reflect differences in the completeness of medical record documentation. The relatively high proportion of stage III in Irkutsk Oblast (22.9 %) alongside a minimal proportion of stage IV (2.1 %) suggests that locally advanced forms are detected before the development of distant metastases.

### Morphological verification, mortality, and cohort of patients under follow-up

According to the state reference book, in 2025 Irkutsk Oblast registered 45 newly diagnosed patients with malignant neoplasms of the eye and adnexa, placing the region 3rd in the Russian Federation after Moscow (93 cases) and Moscow Oblast (75 cases), and ahead of Saint Petersburg (51 cases) [8]. Nationwide, a total of 1,379 patients with pathology under this rubric were registered during the year. In terms of the registered prevalence rate (the cohort of patients under follow-up at the end of the year per 100,000 population), Irkutsk Oblast ranks first among the compared regions—15.6 per 100,000—exceeding the rates of Saint Petersburg (11.9), Moscow (11.7), and Moscow Oblast (7.4). The high prevalence, combined with a moderate absolute number of newly detected cases, reflects favourable survival and the accumulation of the cohort of patients under observation.

The proportion of patients under follow-up for 5 years or more in Irkutsk Oblast was 55.5 %, which is lower than in Moscow (72.1 %) and Saint Petersburg (68.5 %), but comparable to Moscow Oblast (52.8 %). The mortality rate in Irkutsk Oblast proved to be among the lowest among the compared regions—1.9 %, whereas in Moscow it was 2.9 % and in Saint Petersburg 3.0 %; Moscow Oblast registered the lowest value (1.4 %). It should be noted that the reference book text for Irkutsk Oblast also provides an intermediate mortality rate of 2.6 %; the final, refined value adopted for the purposes of this analysis is 1.9 %. The proportion of morphologically verified diagnoses in Irkutsk Oblast was 56.3 %, close to the figure for Moscow (53.7 %), but notably lower than in Moscow Oblast (100 %) and Saint Petersburg (94.3 %). The relatively low frequency of morphological confirmation in UM is a general feature of ophthalmic oncology and is explained by the predominantly clinical-instrumental nature of intraocular tumour diagnosis, in which biopsy carries risks and is not always feasible; the diagnosis is established on the basis of ophthalmoscopy, ultrasound, and optical coherence tomography [1, 5].

The registered prevalence rate warrants separate consideration. The prevalence rate (cohort under follow-up) reflects the cumulative number of living patients and is derived from two parameters—incidence and survival. Consequently, the highest prevalence rate among the compared regions in Irkutsk Oblast (15.6 per 100,000) results from a combination of high incidence and favourable survival: with low mortality and a high proportion of early-stage cases, most newly diagnosed patients accumulate in the follow-up cohort rather than being removed from it by death. Thus, in this instance, the high prevalence of UM is not a sign of unfavourable outcomes but rather a consequence of the simultaneous effect of high incidence and satisfactory treatment results. The proportion of patients under observation for 5 years or more (55.5 %), in turn, depends both on survival and on the duration of the organised registration system; the lower value compared with the capital regions may reflect the relatively recent establishment of the accumulated cohort.

Summarising the obtained results, it should be emphasised that they are internally consistent. The stable absolute number of newly detected cases (37–39 per year), the high proportion of early stages (stage I – 58.3 % in 2025), the low mortality (1.9 %), and the highest registered prevalence among the compared regions (15.6 per 100,000) form a logically coherent picture of a region with simultaneously high incidence and favourable outcomes in the detection and treatment of UM. The territorial analysis demonstrates a concentration of cases in the regional centre while maintaining a significant contribution from rural districts, reflecting both the population distribution and the functioning of the patient referral system to a single specialised facility. Taken together, these characteristics allow Irkutsk Oblast to be regarded as a region with an organised and effective ophthalmic oncology service operating under conditions of elevated incidence. At the same time, the indicator values obtained are based on aggregated data without age standardisation; consequently, they characterise the current epidemiological and organisational situation but do not permit a full differentiation between the contributions of biological risk and detection completeness, which defines the objectives for subsequent in-depth analysis.

## Discussion

The obtained data demonstrate a persistently high incidence of UM in Irkutsk Oblast—approximately 1.6 per 100,000 population per year— which exceeds the average European level by approximately 3–4 times and lies at the upper boundary of values recorded in the most "melanoma-prone" countries of Northern Europe [3, 4]. The region’s leading position in registered prevalence among the constituent entities of the Russian Federation requires a comprehensive explanation that takes into account both biological-geographical and organisational factors.

The first and most likely factor is geographical latitude. Irkutsk Oblast is located at approximately 52° N latitude, corresponding to high latitudes with relatively low annual solar insolation. For UM, in contrast to cutaneous melanoma, an inverse relationship between incidence and solar radiation intensity has been demonstrated: the highest rates are recorded precisely in northern populations—Scandinavia and the northern states [3, 5]. The second factor is the phenotypic composition of the population: among Siberian residents, a significant proportion are individuals with a fair phenotype (fair skin, blue and green eyes), which is an established risk factor for UM development [1, 5]. The combination of high latitude and a "fair" population phenotype creates biological preconditions for elevated incidence, similar to those observed in Northern European countries.

The third factor that must be considered when interpreting inter-regional differences is the probable underdiagnosis of UM in a number of other constituent entities of the Russian Federation. Indirect evidence is provided by the significant proportion of cases with unknown stage in Moscow Oblast (34.7 %) and Saint Petersburg (28.3 %), whereas in Irkutsk Oblast such observations are absent [8]. The presence in the region of an organised ophthalmic oncology service with concentration of patients in a single specialised institution (IROD) contributes to more complete registration and correct staging, which may partially explain both the high prevalence rates and the favourable detection structure. Thus, the high incidence in Irkutsk Oblast likely reflects not only a genuinely increased biological risk but also more complete detectability and registration compared with regions having less developed specialised services.

The possible influence of the age structure of the population should also be considered. UM is predominantly a tumour of older age, with a peak incidence in the 70–79 age group; therefore, differences in the proportion of elderly populations between regions may partially explain differences in crude incidence and prevalence rates. However, the age structure of the population of Irkutsk Oblast, which belongs to regions with a relatively younger population compared with metropolitan agglomerations, makes the high incidence rates even more revealing: if the comparison were performed using standardised rates, the gap between Irkutsk Oblast and other regions would likely be even more pronounced. This consideration indicates the need for age standardisation in subsequent studies.

The favourable characteristics of cancer care in the region deserve special attention. The highest proportion of stage I among the compared regions (58.3 %) and the absence of cases with unknown stage indicate the effectiveness of early diagnosis, which is of fundamental importance for prognosis: timely detection of small tumours allows eye-preserving treatment and is associated with a lower risk of metastasis. According to classical data, the risk of UM metastasis increases proportionally with increasing maximum tumour diameter, which underscores the importance of early-stage detection [2]. One of the lowest mortality rates among the compared regions (1.9 %) and the high accumulated prevalence rate are consistent with the concept of favourable immediate treatment outcomes. At the same time, the relatively low proportion of patients under observation for 5 years or more (55.5 %) compared with the capital regions may reflect both a shorter history of cohort accumulation and specific features of patient referral and registration, and requires further investigation.

Comparison of the obtained data with global epidemiological indicators confirms the high incidence level in Irkutsk Oblast. The obtained rate of approximately 1.6 per 100,000 (i.e. approximately 16 per million) population exceeds the upper limit of the European range (2–8 per million) and even the maximum Scandinavian values (8–10 per million) [3, 4]. However, direct comparison requires caution, because European rates are more often presented as age-standardised coefficients, whereas the present study used a crude rate. Nevertheless, even considering this limitation, the obtained values allow Irkutsk Oblast to be classified among territories with an elevated incidence of UM, comparable to the most "melanoma-prone" regions of Northern Europe, which is biologically consistent given the latitudinal position and phenotypic composition of the population.

Of interest is the comparison of the obtained data with the results of previously conducted domestic studies. According to the Moscow school of ophthalmic oncology, the UM incidence rate in Moscow was approximately 0.9–1.07 per 100,000 adult population [6], which is lower than the rate obtained in the present study for Irkutsk Oblast (≈ 1.6 per 100,000 total population). Studies by S.V. Saakyan and co-authors have also shown that eye-preserving treatment with timely tumour detection provides survival comparable to that with enucleation, confirming the feasibility of early diagnosis and a conservative approach [7]. In this context, the high proportion of stage I in Irkutsk Oblast acquires particular significance, because it is precisely early-stage detection that opens the possibility of using eye-preserving methods with preservation of the eye and visual functions.

The obtained results have practical implications for the organisation of ophthalmic oncology care. The persistently high incidence justifies the need to maintain and develop a specialised ophthalmic oncology service in the region, including ensuring access to modern eye-preserving treatment methods and follow-up observation. The high proportion of early stages and low mortality indicate that the existing system of patient detection and referral in the region functions effectively. At the same time, the relatively low proportion of morphological verification (56.3 %) suggests a potential reserve for improving diagnostic quality through broader implementation of morphological and molecular-genetic confirmation methods, including fine-needle aspiration biopsy in indicated cases.

The present study has a number of limitations. First, the observation period is limited to three years, which does not allow reliable assessment of long-term incidence trends and makes interpretation of interannual fluctuations preliminary. Second, crude (non-standardised) incidence rates without age standardisation were used, which limits the validity of direct comparison with international and other regional data, because differences may be partially attributable to population age structure.

Third, the study did not analyse individual risk factors (phenotype, presence of nevi, tumour molecular-genetic profile) nor detailed treatment outcomes and long-term survival, which is related to the nature of the aggregated source data. Fourth, the stage distribution for 2025 is presented according to the state reference book, whose staging methodology may differ from that used in the regional registry, requiring caution when comparing with data from 2023–2024. These limitations define the directions for further research: conducting age standardisation, extending the observation period, analysing risk factors, and evaluating long-term outcomes of eye-preserving treatment.

Prospects for further research are associated with an in-depth analysis of regional UM epidemiology. The priority task is the calculation of age-standardised incidence rates, which would allow correct comparison of regional data with international and national indicators and eliminate the influence of population age structure. It is advisable to extend the observation period to 10 years or more, as well as to conduct an analysis of long-term survival depending on stage, treatment modality, and tumour localisation. Separate investigation is required for risk factors among the region’s population, including phenotypic characteristics and molecular-genetic features of tumours. The data obtained in the present work may serve as a basis for planning such studies and for optimising the organisation of ophthalmic oncology care in the region.

The obtained data are also relevant in the context of regional public health. Given the persistently high incidence, it appears justified to increase oncological awareness among primary care physicians—polyclinic ophthalmologists and general practitioners—regarding intraocular neoplasms, because timely referral of a patient with suspected choroidal tumour to a specialised facility is a decisive factor for a favourable outcome [1, 2]. Although population-based screening for UM is not performed owing to the rarity of the tumour, opportunistic detection during routine ophthalmological examinations, especially in older age groups with a fair phenotype, can ensure a further increase in the proportion of early stages. The experience of Irkutsk Oblast, characterised by a high proportion of stage I and low mortality, can be regarded as a model of effective organisation of ophthalmic oncology care, reproducible in other regions with similar geographical and population characteristics. The establishment of a regional UM registry with details on localisation, tumour size, treatment methods, and long-term outcomes would allow the accumulated data to be translated into the domain of evidence-based assessment of treatment outcomes and would be a natural extension of the present study.

The identified combination of high latitude of residence and predominance of a fair phenotype in the regional population is consistent with current understanding of the interaction between environmental and hereditary factors in the pathogenesis of UM. In contrast to cutaneous melanoma, for which a direct carcinogenic role of ultraviolet radiation has been proven, in UM the leading role is attributed to constitutional predisposition associated with low melanin content in uveal tract tissues in individuals with light-coloured irides [1, 5]. The northern European populations demonstrating the highest incidence rates are characterised precisely by such phenotypic composition, which makes the analogy with the population of Eastern Siberia biologically plausible. At the same time, the contribution of other, as yet unstudied factors—dietary habits, occupational exposure, and the genetic background of indigenous and migrant populations of the region—cannot be excluded; their elucidation requires specially designed analytical case–control studies. The present descriptive work forms an empirical basis for proposing and subsequently testing such hypotheses, identifying Irkutsk Oblast as a priority territory for in-depth study of UM epidemiology in the Russian Federation.

The interpretation of Irkutsk Oblast’s leading position specifically in registered prevalence deserves particular comment. Prevalence (the cohort of patients under follow-up) is an accumulated indicator over many years, depending both on the incidence level and on survival and the quality of dispensary registration [8]. The region’s first place in this indicator (15.6 per 100,000) alongside high incidence and low mortality is logically explainable: patients detected at early stages and successfully treated with eye-preserving methods remain under observation for long periods, forming a substantial accumulated cohort.

Conversely, in regions with high mortality or incomplete dispensary registration, cohort accumulation occurs more slowly. Thus, Irkutsk Oblast’s leadership in prevalence should be interpreted not as an indicator of unfavourable outcomes but as an integral result of the combination of high incidence with effective organisation of detection and follow-up. This fundamental distinction between incidence and prevalence indicators is important to consider when formulating management decisions, because direct comparison of regions without differentiating these categories may lead to erroneous conclusions about the quality of cancer care.

## Conclusions

1. Over the period 2023–2025, 112 new cases of uveal melanoma were registered in Irkutsk Oblast (37, 36, and 39 by year), with a stable crude incidence rate of approximately 1.6 per 100,000 population per year, which substantially exceeds the average European level and previously published data for Moscow.
2. In 2025, Irkutsk Oblast ranked 3rd in the Russian Federation in the number of newly registered patients with neoplasms of the eye and adnexa (45 individuals) and 1st in registered prevalence (15.6 per 100,000 population), surpassing Moscow, Moscow Oblast, and Saint Petersburg.
3. The region is characterised by a favourable detection structure: the highest proportion of stage I among the compared regions (58.3 %), the absence of cases with unknown stage, and one of the lowest mortality rates (1.9 %), indicating the effectiveness of early diagnosis and the organisation of specialised ophthalmic oncology care.
4. The high incidence of uveal melanoma in Irkutsk Oblast is likely attributable to a combination of high geographical latitude, the predominance of a fair phenotype among the Siberian population, and more complete detectability in the context of an organised service; further studies with age standardisation of indicators and extension of the observation period are required to clarify the true level of risk.

## Data Availability

All data produced in the present work are contained in the manuscript

